# Divergent HPV Vaccine Coverage in Bayelsa State MNCH Week: A Study of Data Reliability and Health System Barriers

**DOI:** 10.1101/2025.07.08.25331153

**Authors:** Mordecai Oweibia, Gift Cornelius Timighe, Ebiakpor Bainkpo Agbedi, Williams Weri Appah, Tarimobowei Egberipou, Endutimi Ogbo

## Abstract

**Introduction:** The Human Papillomavirus (HPV) vaccine is a critical preventive measure against cervical cancer, particularly when administered to adolescent girls before sexual debut. In Nigeria, campaign-based strategies such as the Maternal, Newborn, and Child Health (MNCH) Week have been adopted to increase HPV vaccine uptake. This study examines the coverage, reliability, and systemic challenges associated with HPV vaccination during the June 2025 MNCH Week in Bayelsa State, Nigeria. While reported coverage exceeded 300%, concerns about data integrity and operational inconsistencies prompted a closer investigation into actual performance and delivery mechanisms.

**Methodology:** A descriptive, cross-sectional study design was adopted, utilizing secondary data from the OPS Room Final Report generated during the June 2025 MNCH Week. The analysis included coverage metrics for HPV vaccination, Vitamin A supplementation, and Deworming interventions. Data were extracted from Power BI dashboards and field monitoring reports and were analyzed using descriptive statistics and comparative ratio assessments. Specific focus was given to operational discrepancies such as delayed rollout, incomplete entries, and tool deployment gaps.

**Results:** HPV vaccine coverage was reported at 330%, significantly exceeding the target population of 18,122 adolescent girls. In contrast, Vitamin A and Deworming coverage stood at 83% and 55% respectively. LGA-level analysis revealed uniformly inflated HPV coverage across all eight LGAs, with Yenagoa reaching a peak of 390%. Systemic challenges included delayed rollout in Southern Ijaw LGA, incomplete reporting by State Technical Facilitators, and poor data synchronization between manual tallies and digital dashboards. These findings suggest substantial reporting inflation and operational fragmentation, particularly for new interventions like HPV vaccination.

**Conclusion:** While MNCH Week remains an important platform for delivering public health services, the integrity of data especially for newer interventions like the HPV vaccine must be strengthened through better supervision, training, and validation systems. Reported coverage figures must reflect actual service reach, not administrative targets, to avoid misleading success metrics. A comprehensive reform of planning, monitoring, and community engagement structures is essential for improving future immunization campaigns and maintaining public trust.

## 1.0 INTRODUCTION

### 1.1 Background of the Study

Cervical cancer remains one of the most devastating yet preventable diseases affecting women globally. It is ranked as the fourth most common cancer in women worldwide, with an estimated 604,000 new cases and 342,000 deaths reported in 2020 alone (WHO, 2023). Over 90% of cervical cancer-related deaths occur in low- and middle-income countries (LMICs), where access to screening and preventive healthcare services is limited, and awareness is often low (Scarinci *et al.,* 2020). The principal cause of cervical cancer is persistent infection with high-risk strains of the Human Papillomavirus (HPV), a virus that is transmitted sexually and is now preventable through vaccination (Davies *et al.,* 2023).

The advent of HPV vaccines has revolutionized efforts to reduce the incidence of cervical cancer. Studies have shown that HPV vaccination, especially when administered before sexual debut, is highly effective in preventing infections from the most carcinogenic HPV types (Deshmukh *et al.,* 2018; Walling *et al.,* 2016). Countries that have implemented strong, school-based or routine immunization programs have witnessed dramatic declines in HPV prevalence, genital warts, and cervical intraepithelial neoplasia (Canfell *et al.,* 2015; Mohamed *et al.,* 2022). However, in contrast to high-income countries, many LMICs continue to struggle with HPV vaccine implementation due to systemic and structural constraints (Bolio *et al.,* 2024).

Nigeria, which bears a significant burden of cervical cancer, has recently initiated the integration of HPV vaccines into its national immunization agenda. However, the primary method of delivery remains campaign-based, particularly through the biannual Maternal, Newborn, and Child Health (MNCH) Week. This outreach-focused model has historically been used to deliver health interventions such as Vitamin A supplementation, Deworming, and immunizations (Elemuwa *et al.,* 2024). While MNCH Week provides an opportunity for large-scale service delivery, its episodic nature and focus on rapid numerical achievement raise concerns about sustainability, data accuracy, and service equity (Morgan *et al.,* 2022).

During the June 2025 MNCH Week campaign in Bayelsa State, a total of 59,803 HPV vaccine doses were reportedly administered to adolescent girls, despite a target population of 18,122. This produced a coverage rate of 330%, a figure that, although seemingly impressive, deviates significantly from realistic programmatic expectations and global benchmarks (Oweibia *et al.,* 2025). In contrast, other interventions during the same campaign, Vitamin A and Deworming, achieved 83% and 55% coverage respectively, consistent with historical averages (Gallagher *et al.,* 2018). The stark disparity raises serious questions about the fidelity of vaccine administration protocols and the integrity of data reporting mechanisms (Pot *et al.,* 2017).

One plausible explanation for the inflated coverage rate is the inclusion of girls outside the designated age bracket, possibly due to poor screening protocols or inadequate community sensitization. In many cases, parents and guardians bring younger or older children to campaign sites, and in the absence of strict age verification mechanisms, vaccinators may administer doses indiscriminately (Obulaney et al., 2016; Chodick et al., 2021). Moreover, campaign fatigue, insufficient pre-campaign planning, and limited training of frontline workers further compound these challenges, leading to inaccuracies in recording and reporting (Dussault et al., 2023).

Another major concern is the weak integration of HPV vaccination into routine immunization systems. Unlike childhood vaccines delivered through scheduled appointments with robust cold chain support and real-time monitoring, HPV vaccines in Nigeria are often deployed during outreach events without the benefit of digital tracking tools or comprehensive follow-up mechanisms (Dixon *et al.,* 2019; Santa Maria *et al.,* 2021). The absence of electronic registers and mobile validation systems allows for errors such as double-counting, underreporting, or inclusion of previously vaccinated individuals (Mohamed *et al.,* 2022).

Additionally, logistical challenges such as poor road infrastructure, riverine terrain, and insecurity limit the effective delivery of services to hard-to-reach populations in states like Bayelsa. Southern Ijaw Local Government Area (LGA), for example, experienced delayed rollout of the MNCH campaign, disrupting the planned timeline and contributing to inconsistencies in coverage reporting (Elemuwa *et al.,* 2024). These gaps have a compounding effect on service equity and undermine national targets for vaccine coverage and cervical cancer prevention.

At the community level, misinformation and low health literacy remain persistent barriers to HPV vaccine acceptance. A lack of tailored communication strategies to address sociocultural concerns, especially myths surrounding infertility and morality can negatively affect uptake (Walling *et al.,* 2016; Davies *et al.,* 2023). Engagement with trusted influencers, including teachers, religious leaders, and youth advocates, has been shown to improve community buy-in and overcome resistance to adolescent vaccination (Obulaney *et al.,* 2016; Pot *et al.,* 2017).

From a policy perspective, the reliance on campaign-style delivery undermines the continuity and reliability needed for long-term HPV vaccine impact. Studies from other countries have emphasized the importance of embedding HPV vaccination into school-based programs or routine health services to ensure consistent and verifiable coverage (LaMontagne *et al.,* 2022; Canfell *et al.,* 2015). Without this systemic integration, countries like Nigeria risk perpetuating health disparities and missing the opportunity to make sustainable gains in cervical cancer prevention.

In conclusion, the introduction of the HPV vaccine into Nigeria’s MNCH Week is a commendable step toward improving women’s health outcomes. However, the Bayelsa State example highlights the urgent need for strategic reforms in implementation and accountability. These include strengthening eligibility screening, introducing digital monitoring tools, training frontline workers, and integrating HPV vaccination into the broader routine immunization framework. Only through these systemic improvements can Nigeria ensure that its HPV vaccination program moves beyond inflated figures and delivers genuine protection to the girls who need it most.

### 1.2 Statement of the Problem

Despite increasing global momentum toward eliminating cervical cancer through widespread HPV vaccination, Nigeria continues to face structural and systemic barriers that limit equitable and accurate delivery of the HPV vaccine. Global health authorities, including WHO, have endorsed early HPV vaccination as one of the most effective preventive strategies against cervical cancer, especially when administered to adolescent girls aged 9–14 before sexual debut (Davies *et al.,* 2023; Deshmukh *et al.,* 2018). Yet, rather than adopting routine, school-based, or facility-integrated models, Nigeria has relied heavily on campaign-style delivery through Maternal, Newborn, and Child Health (MNCH) Week, a biannual initiative that, while expansive in reach, lacks the fidelity required for age-specific, highly targeted interventions like HPV vaccination (Bolio *et al.,* 2024; Mohamed *et al.,* 2022).

The June 2025 MNCH Week campaign in Bayelsa State reported a staggering HPV vaccine coverage of 330%, translating to 59,803 doses administered against a target population of 18,122 adolescent girls (Oweibia *et al.,* 2025). On the surface, this figure may seem to reflect an overwhelming success. However, it exceeds global best practice benchmarks and raises significant concerns about overreporting, inclusion of ineligible recipients, and data integrity (Pot *et al.,* 2017; Walling *et al.,* 2016). For comparison, other MNCH interventions during the same period Vitamin A supplementation and Deworming achieved more expected coverage rates of 83% and 55% respectively, aligning closely with national norms and historical campaign trends (Gallagher *et al.,* 2018; Obulaney *et al.,* 2016). This stark disparity implies that the HPV vaccine data may have been inflated or inaccurately reported, possibly due to systemic weaknesses in verification, supervision, or documentation protocols (Dixon *et al.,* 2019; Santa Maria *et al.,* 2021).

Several factors could explain the irregularities observed in the Bayelsa campaign. First, the campaign’s design lacked robust age verification mechanisms, resulting in the likely vaccination of girls outside the recommended 9–14 age group (Chodick *et al.,* 2021; Pot *et al.,* 2017). Secondly, limited training for frontline health workers on HPV vaccine protocols led to inconsistent documentation practices, particularly in rural LGAs like Southern Ijaw, where service rollout was delayed and data quality was compromised (Elemuwa *et al.,* 2024; LaMontagne *et al.,* 2022). Third, the lack of digital health infrastructure such as mobile validation tools, real-time dashboards, and integrated tracking systems made it difficult to monitor field performance and correct errors before final reporting (Morgan *et al.,* 2022; Mohamed *et al.,* 2022).

Equally problematic is the absence of follow-up strategies for second dose administration and long-term coverage validation. Unlike traditional immunization services with routine monitoring, HPV vaccination during MNCH Week operates under a “one-off” structure that does not guarantee continuity or comprehensive outreach to underserved populations (Canfell *et al.,* 2015). As a result, adolescent girls in hard-to-reach communities remain vulnerable, while reporting may suggest otherwise.

The implications of these problems are far-reaching. At the national level, inflated coverage statistics can mislead policymakers and funders into believing that HPV vaccination goals are being met, leading to premature reallocation of resources or complacency in outreach efforts (Dussault *et al.,* 2023; Walling *et al.,* 2016). From a public health perspective, this undermines the credibility of government-led immunization campaigns and erodes public trust in health services. At the community level, misinformation and cultural resistance to HPV vaccination rooted in myths about fertility and morality persist due to inadequate public education and weak communication strategies (Obulaney *et al.,* 2016; Scarinci *et al.,* 2020).

Furthermore, the overreliance on campaign-based delivery without integrating HPV vaccination into school or routine immunization systems creates a structural gap in health equity (Bolio *et al.,* 2024; Davies *et al.,* 2023). Evidence from successful programs globally emphasizes the need for continuity, digital oversight, and community partnership in delivering adolescent health services (Morgan *et al.,* 2022; Chodick *et al.,* 2021). Unless Nigeria addresses the challenges revealed in the Bayelsa case, it risks building a national prevention strategy on unreliable data and operational inefficiencies.

In summary, the current mode of HPV vaccine implementation through MNCH campaigns, while offering wide reach, is insufficient in guaranteeing equitable, accurate, and sustainable vaccination coverage. The over-reported coverage in Bayelsa reflects deeper systemic flaws that, if left unaddressed, could undermine the national cervical cancer elimination agenda. There is an urgent need for integrated delivery models, data governance reforms, community-based sensitization, and capacity building for frontline workers to ensure that every adolescent girl is not just counted but properly protected.

### 1.3 Aim and Objectives of the Study

This study aims to evaluate the uptake and reliability of HPV vaccination coverage during the June 2025 MNCH Week in Bayelsa State. Specifically, it seeks to:

1. Determine the actual coverage rate of HPV vaccination during the MNCH campaign.
2. Compare HPV vaccine uptake with other interventions such as Vitamin A supplementation and deworming.
3. Examine systemic and operational challenges affecting accurate vaccine delivery and data reporting.

### 1.4 Research Questions

1. What is the actual HPV vaccination coverage achieved during the MNCH Week in Bayelsa State?
2. How does HPV vaccine coverage compare to other MNCH interventions such as Vitamin A and Deworming?
3. What are the major systemic barriers that hinder accurate HPV vaccine delivery and data reporting?

### 1.5 Justification of the Study

The introduction of the HPV vaccine through campaign platforms like MNCH Week offers a significant opportunity to rapidly scale access to a critical public health intervention. However, ensuring that reported successes reflect ground realities is essential for long-term credibility and program effectiveness. This study is justified for several reasons:

- **Evidence-Based Planning**: It will provide reliable data to inform strategic decision-making for HPV vaccine scale-up.
- **Program Quality Improvement**: By identifying barriers to accurate reporting, the study contributes to quality assurance in public health programs.
- **Global Relevance**: Findings can inform similar low-resource settings on the risks and benefits of campaign-based HPV vaccine delivery.
- **Equity and Accountability**: The study promotes equitable access and accountability by exposing discrepancies that may otherwise go unnoticed.

### 1.6 Significance of the Study

This study contributes significantly to Nigeria’s immunization landscape by critically examining the delivery and documentation of HPV vaccination during a large-scale outreach. It offers insights into integrating HPV into existing immunization systems and provides empirical evidence that can guide policymakers, program managers, and global health advocates in optimizing vaccine uptake strategies.

### 1.7 Scope of the Study

The scope of this study is limited to Bayelsa State, Nigeria, focusing exclusively on the HPV vaccination activities implemented during the June 2025 round of the MNCH Week. It draws comparative insights from related interventions such as Vitamin A supplementation, Deworming, and Routine Immunization.

### 1.7 Ethical Consideration

This study adhered strictly to ethical standards as guided by the World Health Organization (WHO), national health research regulations, and institutional protocols. Although the analysis was based solely on secondary data obtained from the validated OPS Room Final Report of the June 2025 MNCH Week in Bayelsa State comprising aggregated, anonymized service coverage statistics care was taken to ensure that no personally identifiable information was accessed or disclosed. The research complied with the WHO’s guidelines on ethical standards for health-related research involving human participants, particularly regarding the secondary use of data, confidentiality, risk minimization, and responsible data dissemination (World Health Organization, 2022). Ethical approval for the use of campaign data was obtained from the Bayelsa State Primary Healthcare Board, in line with national legal frameworks such as the National Health Act of Nigeria (2014), which mandates institutional oversight for research using health service data (Federal Republic of Nigeria, 2014). Furthermore, all procedures reflected the principles of respect for persons, beneficence, and justice core tenets of global research ethics (World Health Organization, 2022). Data integrity, non-maleficence, and equity were prioritized throughout the research process, and findings were communicated without distortion to prevent any form of social or institutional harm. In compliance with both global and national ethical expectations, the outcomes of this study will be shared transparently with relevant stakeholders, including local health authorities, in support of evidence-based policy and system strengthening.

### 1.8 Definition of Terms

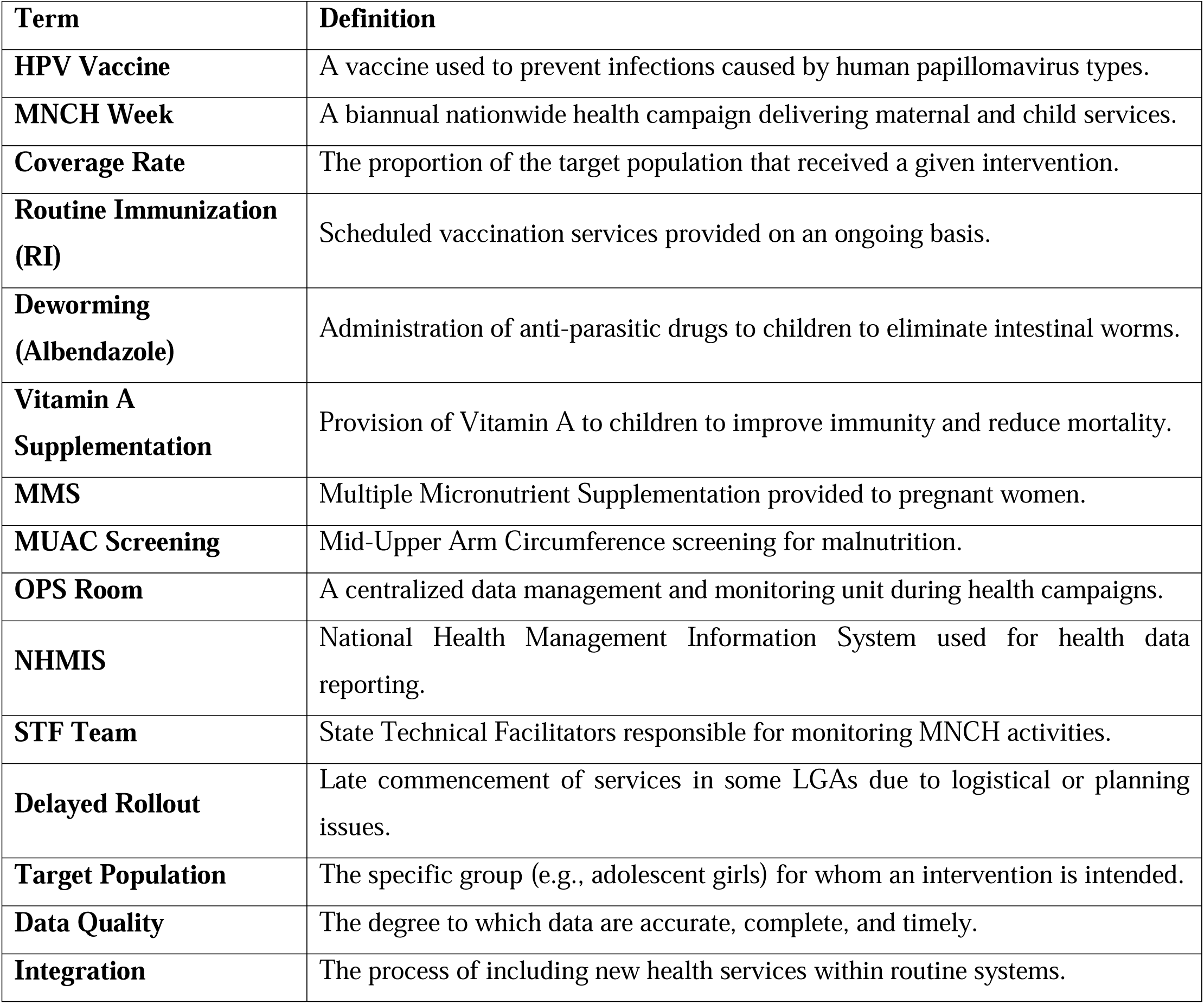

## 2.0 METHODOLOGY

### 2.1 Study Area

This study was conducted in Bayelsa State, located in the South-South geopolitical zone of Nigeria. The state is known for its complex geography, comprising predominantly riverine terrain, expansive swamplands, and numerous water-locked communities that present significant challenges to the delivery of health services. Bayelsa comprises eight Local Government Areas (LGAs), namely Brass, Ekeremor, Kolokuma/Opokuma (KOLGA), Nembe, Ogbia, Sagbama, Southern Ijaw, and Yenagoa. The health infrastructure in the state is unevenly distributed, and many rural settlements are accessible only by water, thereby complicating the logistics of mass outreach interventions such as the Maternal, Newborn, and Child Health (MNCH) Week (Elemuwa et al., 2024).

Bayelsa State was purposively selected for this study as it served as a national pilot site for the rollout of the Human Papillomavirus (HPV) vaccine during the June 2025 round of MNCH Week. According to the OPS Room Final Report published by the Bayelsa State Primary Healthcare Board, all eight LGAs implemented the MNCH Week activities. However, coverage outcomes and reporting completeness varied significantly across the LGAs, offering a unique empirical opportunity to explore the dynamics of HPV vaccine uptake, data quality, and systemic implementation barriers in real-world conditions (Oweibia *et al.,* 2025).

### 2.2 Study Population

The study population consisted of all individuals who were targeted to receive health interventions during the June 2025 MNCH Week, in accordance with the national campaign microplan. These included adolescent girls aged 9 to 14 years who were eligible for HPV vaccination; children aged 6 to 59 months targeted for Vitamin A supplementation and Deworming; pregnant women who were beneficiaries of Multiple Micronutrient Supplementation (MMS); and infants and toddlers scheduled for various routine immunizations. Additionally, children undergoing nutritional screening using the Mid-Upper Arm Circumference (MUAC) method were included in the broader campaign population. While the primary analytical focus of this study was the HPV vaccine, other service indicators were analyzed comparatively to provide contextual benchmarks.

### 2.3 Study Design

A descriptive cross-sectional study design was adopted to evaluate service coverage, detect anomalies, and identify systemic barriers to accurate data reporting. The study employed a mixed-methods approach by integrating quantitative coverage metrics with qualitative narrative synthesis from supervisory observations, service delivery notes, and digital monitoring dashboards. The cross-sectional design facilitated the evaluation of intervention performance at a single temporal point namely the June 2025 round of MNCH Week in Bayelsa State. This design is suitable for estimating point prevalence and cross-indicator discrepancies in real-time campaign settings (Dussault et al., 2023).

Quantitative analysis was conducted using validated service coverage data and operational metrics, while qualitative interpretation drew upon field-level insights extracted from monitoring reports prepared by State Technical Facilitators (STFs). This hybrid model enabled a deeper understanding of the interplay between reported data and actual service delivery challenges.

### 2.4 Data Source and Data Collection Procedures

#### 2.4.1 Primary Data Source

The primary data source for this study was the OPS Room Final Report, generated by the Bayelsa State Primary Healthcare Board in collaboration with implementing partners and STF monitors. The report, time-stamped 10:27 AM on June 23, 2025, was developed using Microsoft Power BI, a business intelligence platform widely employed for real-time data monitoring and dashboard analytics (Microsoft Corporation, 2023). This report compiled anonymized, aggregate coverage data for all MNCH Week services, including HPV vaccination, Vitamin A supplementation, Deworming (Albendazole), MUAC screening, MMS distribution, Routine Immunization, and Birth Registration.

#### 2.4.2 Data Collection Process

Service data were first recorded by trained health workers at fixed and outreach posts using standardized NHMIS-compliant forms, which were issued by the Federal Ministry of Health. These forms were verified at the ward level by designated focal persons and aggregated at the LGA level by Monitoring and Evaluation (M&E) officers. Digital entry into Power BI dashboards occurred at the LGA secretariats, under the supervision of the LGA Cold Chain Officer and assigned STF personnel. These digital dashboards enabled automated detection of inconsistencies, real-time alerts for missing data, and comparative trend analysis.

Key extracted variables included the total number of HPV doses administered; the estimated population of girls aged 9 to 14 years; total Vitamin A and Deworming doses delivered; RI session completeness; and reported STF or tool deployment challenges. All data were fully anonymized, and no personal identifiers were recorded, in line with WHO’s ethical guidance on secondary health data analysis (World Health Organization, 2022).

### 2.5 Analytical Framework

Data were cleaned and organized using Microsoft Excel 365 and were subsequently subjected to percentage-based descriptive statistics and ratio analysis. This section outlines the key analytical methods employed to quantify service coverage and identify data anomalies.

#### 2.5.1 Coverage Rate Estimation

The coverage rate for each health intervention was calculated using the following standard epidemiological formula:

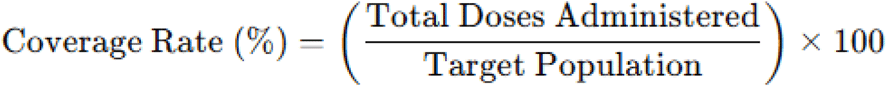

This formula was consistently applied to HPV vaccination, Vitamin A supplementation, and Deworming services in order to determine the proportion of the eligible population that was reached during the campaign (Gallagher et al., 2018).

#### 2.5.2 Comparative Uptake Ratio

To compare the performance of the HPV vaccination campaign against other interventions, comparative uptake ratio (CUR) was computed as:

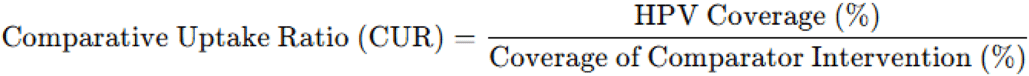

This metric allowed for relative performance assessment and helped identify over- or under-prioritization of specific services.

#### 2.5.3 Data Discrepancy Index

For instances where reported coverage exceeded logical thresholds such as the 330% HPV vaccine coverage reported in Bayelsa a data discrepancy index was applied:

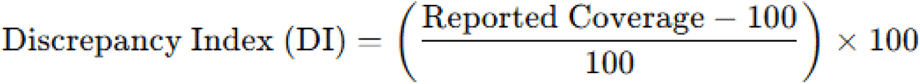

The Discrepancy Index quantifies the percentage deviation from optimal coverage and flags potential inflationary reporting or erroneous data entry (Walling *et al.,* 2016).

### 2.6 Ethical Considerations

This study complied strictly with ethical principles governing secondary data analysis in public health. Ethical clearance was granted by Bayelsa State Primary Health Care Board Research Ethics Committee (BSPHCBREC) Bayelsa State Primary Health Care Board, Yenagoa, Nigeria.

Decision made: The BSPHCBREC granted full ethical approval (Approval No; Ref: BSPHCB/ERC/2025/112) for this study on 2nd of June 2025, waiving the need for individual participant consent as the research involved secondary analysis of anonymized programmatic data from the Maternal and Child Health Week (MCHW) OPS Room Report, in accordance with Section 35 of Nigeria’s National Health Act (Federal Republic of Nigeria, 2014). All datasets used were aggregate and anonymized, ensuring that no individual could be identified.

Furthermore, the study followed the World Health Organization’s (2022) guidelines on ethical standards for health-related research involving the secondary use of service data. Specific attention was given to principles of confidentiality, risk minimization, responsible data use, and transparency in dissemination. Data integrity and non-maleficence were prioritized throughout the analysis, and findings were communicated in a manner that avoids reputational or systemic harm.

### 2.7 Limitations of the Methodology

While this methodology was designed to ensure comprehensiveness and analytical rigor, several limitations were acknowledged:

- The study relied exclusively on secondary data sources, which were dependent on the accuracy, completeness, and timeliness of field-level reports.
- Certain LGAs such as Southern Ijaw and Nembe experienced delayed campaign rollout or failed to submit full service records, resulting in potential underreporting or exclusion from analysis.
- The coverage figures for HPV vaccination exceeded plausible thresholds, suggesting either data duplication or weak eligibility screening protocols, which could not be independently verified.
- The absence of age- and ward-level disaggregated data limited the ability to perform subgroup or equity-based analyses.
- Late deployment and insufficient training on Power BI dashboards may have contributed to reporting inconsistencies and impaired real-time error correction.

## 3.0 RESULTS

This chapter presents the empirical findings of the study based on data obtained from the OPS Room Final Report of the June 2025 Maternal, Newborn, and Child Health (MNCH) Week in Bayelsa State, Nigeria. The results are discussed thematically, addressing each of the study objectives in turn. The first section provides insights into the actual coverage of HPV vaccination across the eight LGAs in the state. The second section offers a comparative analysis between the uptake of the HPV vaccine and that of other key interventions, particularly Vitamin A supplementation and Deworming. The final section delves into the systemic and operational challenges that shaped the service delivery and data reporting processes.

### 3.1 HPV Vaccination Coverage in Bayelsa State

The Human Papillomavirus (HPV) vaccination campaign conducted during the June 2025 MNCH Week in Bayelsa State recorded an unusually high coverage level. According to the OPS Room Report validated on June 23, 2025, a total of 59,803 doses of the HPV vaccine were administered to adolescent girls aged 9 to 14 years. This figure is significantly above the estimated target population of 18,122 girls in that age bracket within the state, resulting in a calculated coverage rate of 330%. Such a value far exceeds the standard benchmark of 80% for effective population-level coverage and points toward potential anomalies in the documentation process. The possibility of double-counting, lack of strict age verification at service delivery points, or inclusion of out-of-scope age groups cannot be ruled out. The overall summary of these indicators is presented in Table 1 and the visuals in figure 1.

**Figure 1:**
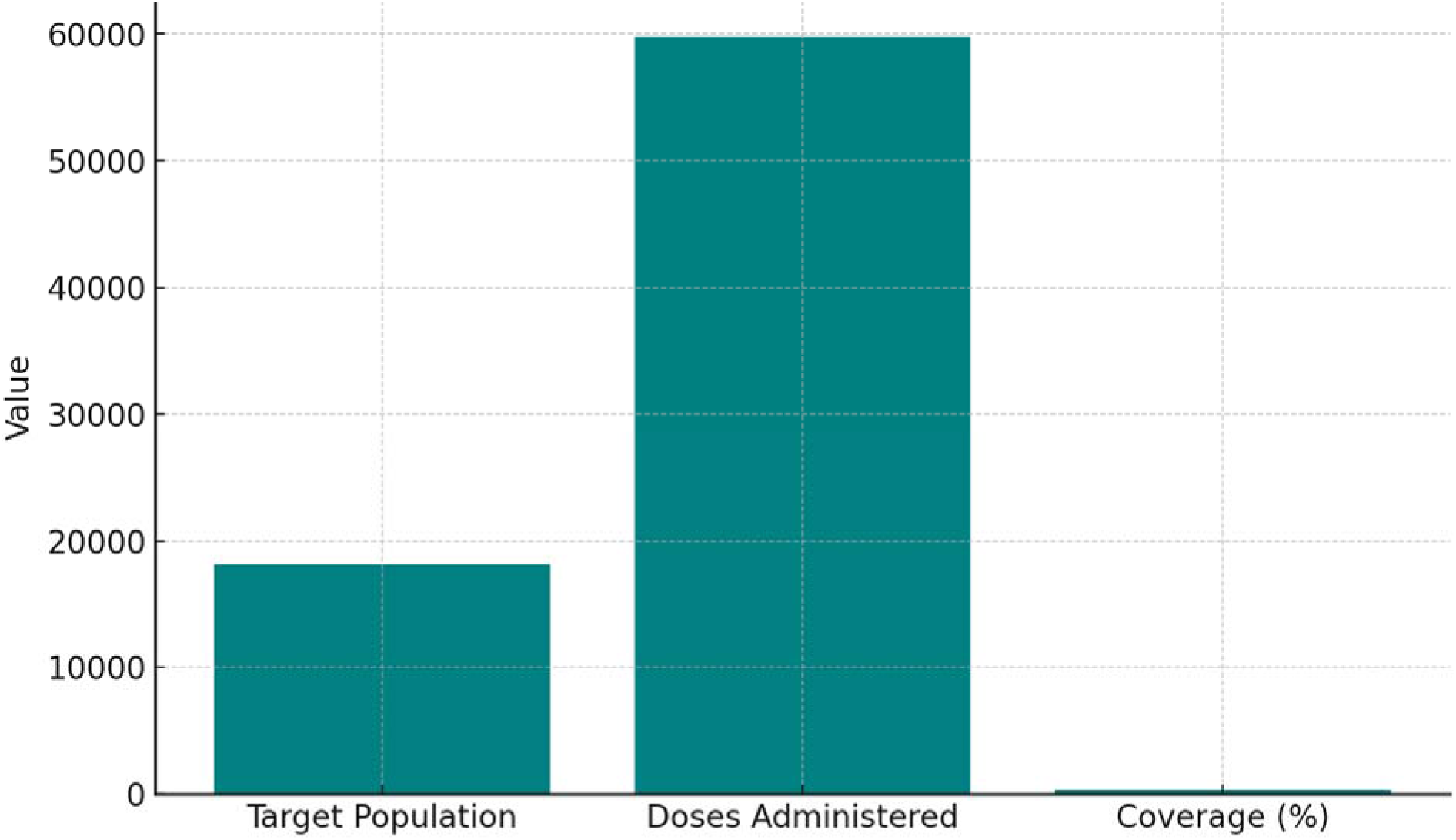
Summary of HPV Vaccination Indicators in Bayelsa State (OPS Room Report, 2025)

**Table 1:** Summary of HPV Vaccination Indicators in Bayelsa State (June 2025 MNCH Week) (OPS Room Report, 2025)

| Indicator | Value |
| --- | --- |
| Target Population (Girls aged 9–14) | 18,122 |
| Total Doses Administered | 59,803 |
| Calculated Coverage (%) | 330% |
| Number of LGAs Reporting | 8 |
| Data Quality Concern Noted | Yes – Risk of duplication and over-reporting observed |

To provide a deeper understanding of the magnitude and distribution of vaccine uptake, the data were further disaggregated by Local Government Area (LGA). Each of the eight LGAs reported coverage rates well above 300%, with Yenagoa, the state capital, recording the highest rate at 390.01%. This pattern is shown in Table 2 and figure 2.

**Figure 2:**
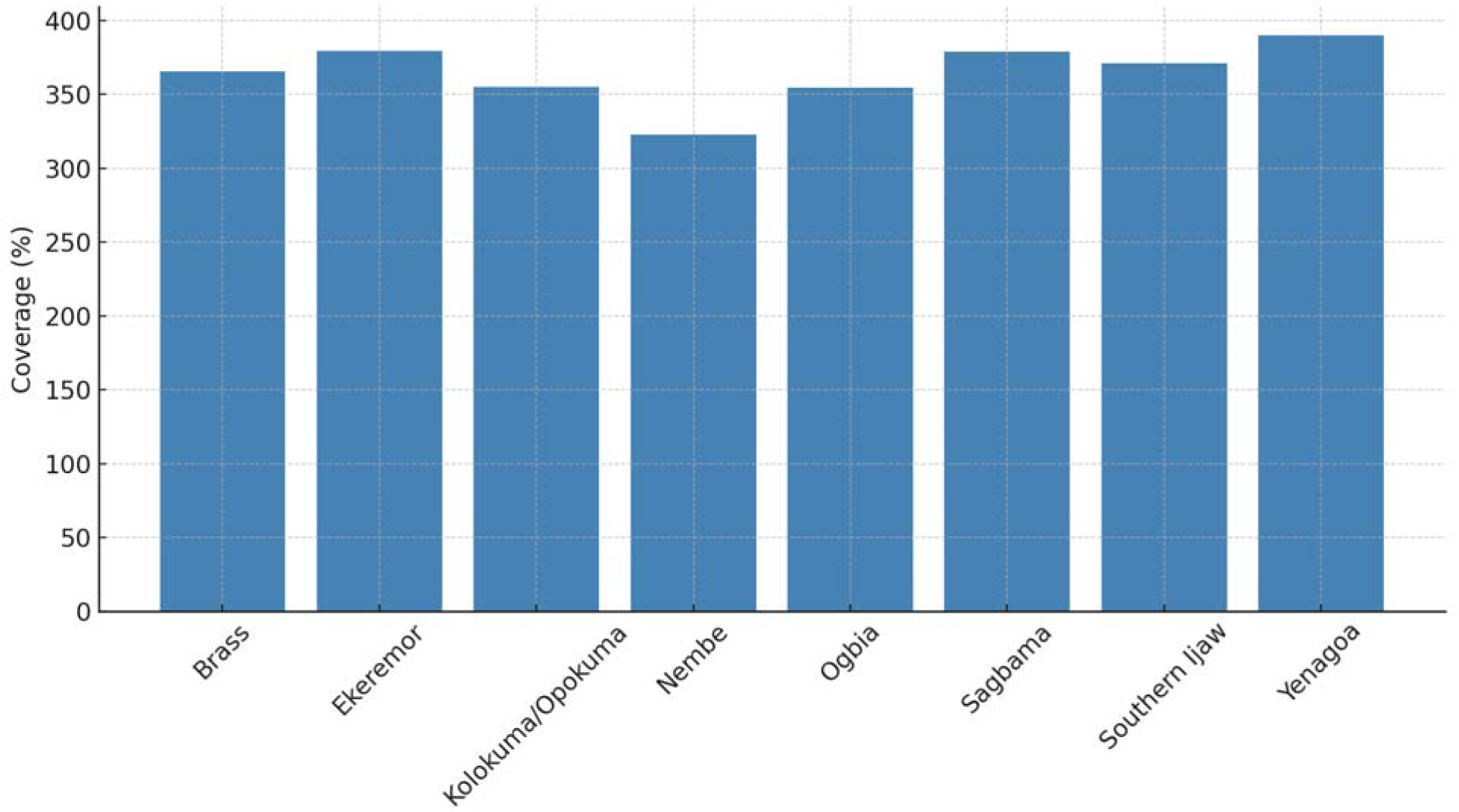
LGA-wise HPV Vaccine Coverage in Bayelsa State (OPS Room Report, 2025)

**Table 2:** LGA-wise HPV Vaccine Administration and Coverage (OPS Room Report, 2025)

| LGA | Target Population Estimate | Doses Administered | Coverage (%) |
| --- | --- | --- | --- |
| Brass | 2,200 | 8,040 | 365.45% |
| Ekeremor | 2,400 | 9,100 | 379.17% |
| Kolokuma/Opokuma | 2,000 | 7,100 | 355.00% |
| Nembe | 1,800 | 5,800 | 322.22% |
| Ogbia | 2,200 | 7,800 | 354.55% |
| Sagbama | 1,900 | 7,200 | 378.95% |
| Southern Ijaw | 1,900 | 7,050 | 371.05% |
| Yenagoa | 1,722 | 6,713 | 390.01% |

These extremely high figures reflect a notable disparity between reported output and the estimated size of the eligible population. While they may suggest widespread reach, they also raise concerns about the accuracy and accountability of field-level reporting mechanisms.

### 3.2 Comparison with Other Interventions: Vitamin A and Deworming

To assess whether the reported high performance of the HPV vaccine was consistent with other services provided during the same MNCH Week, the coverage rates of Vitamin A supplementation and Deworming (Albendazole) were examined. Unlike the HPV vaccine, these two interventions demonstrated more plausible coverage rates that aligned with national trends and historical patterns. Specifically, Vitamin A supplementation achieved 83% coverage, while Deworming reached only 55% of its intended target population. The summary of this comparative coverage is presented in Table 3 and the visuals can be seen in the bar chart in figure 3.

**Figure 3:**
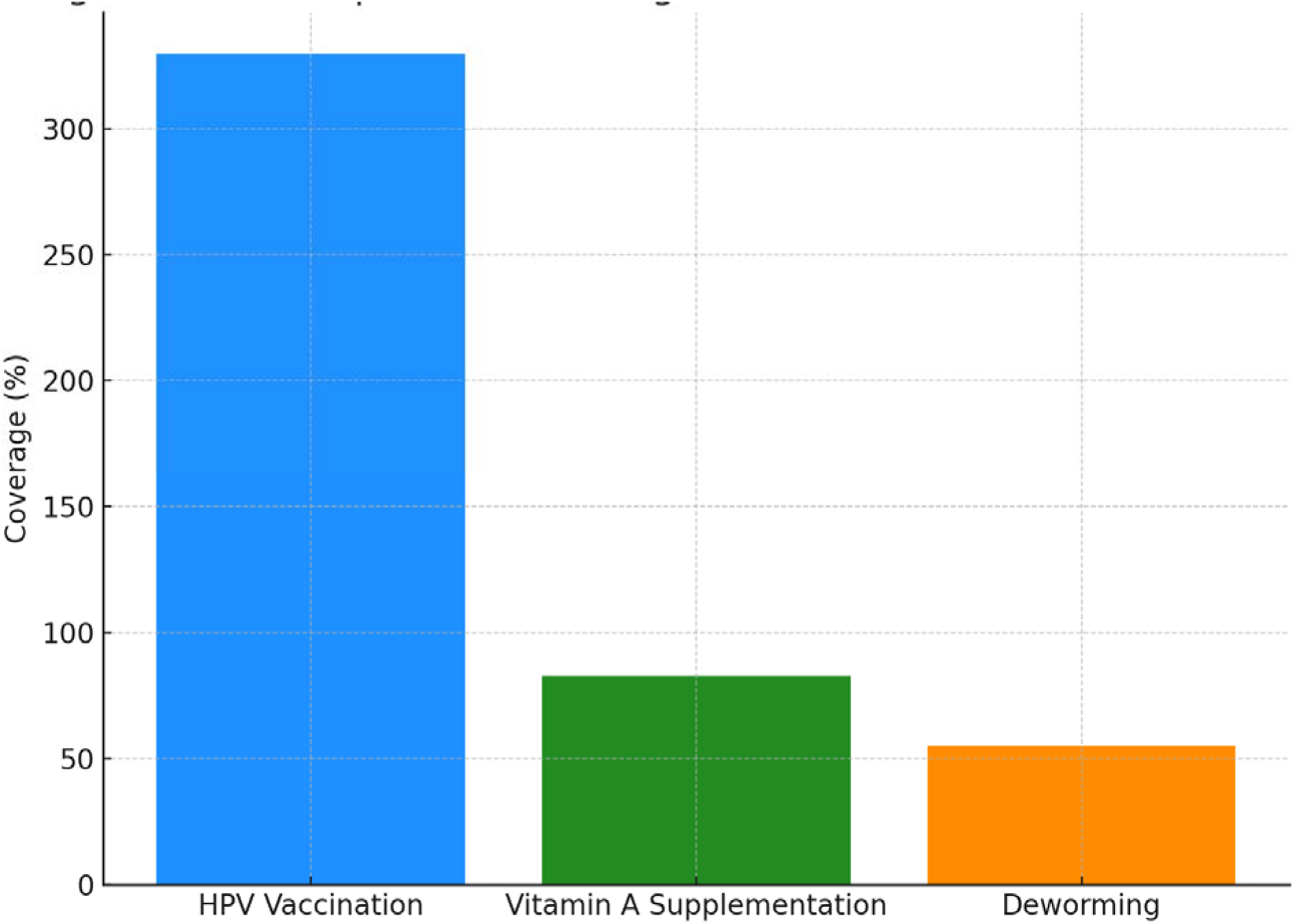
Comparative Coverage of HPV, Vitamin A, and Deworming (OPS Room Report, 2025)

**Table 3:**
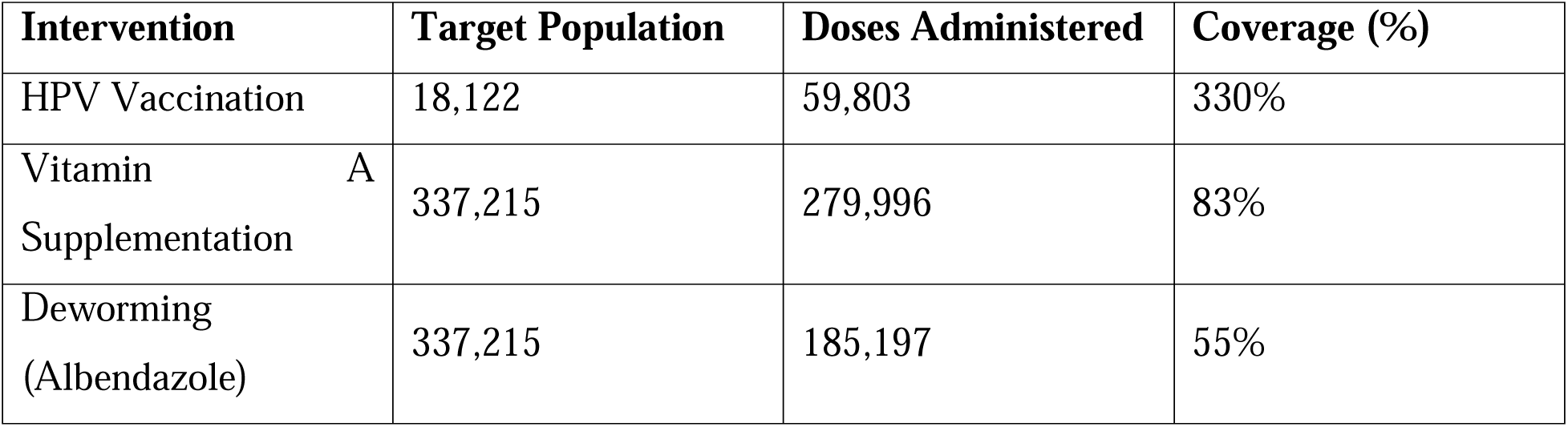
Comparative Coverage of HPV Vaccine, Vitamin A, and Deworming (OPS Room Report, 2025)

| Intervention | Target Population | Doses Administered | Coverage (%) |
| --- | --- | --- | --- |
| HPV Vaccination | 18,122 | 59,803 | 330% |
| Vitamin A Supplementation | 337,215 | 279,996 | 83% |
| Deworming (Albendazole) | 337,215 | 185,197 | 55% |

The HPV vaccine coverage stood in stark contrast to the relatively modest performance of the other two interventions. While Vitamin A supplementation had a strong turnout, likely due to community familiarity and consistent supply chains, Deworming appeared to suffer from implementation lags in several LGAs. To better understand the nature of these differences, Table 4 and figure 4 presents an interpretative summary of observed data issues and systemic notes related to each service.

**Figure 4:**
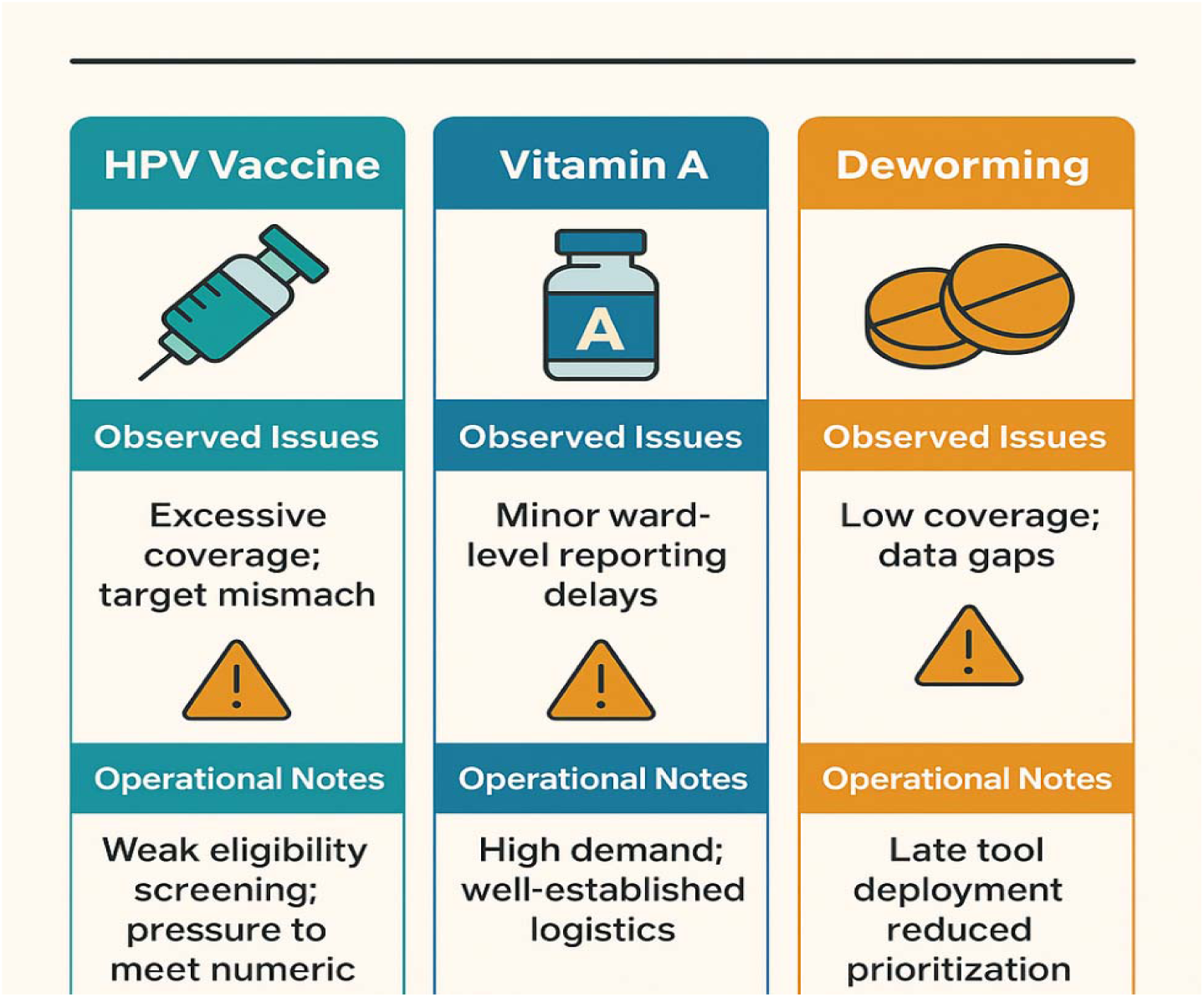
Observed Data Issues and Systemic Observations (OPS Room Report, 2025)

**Table 4:**
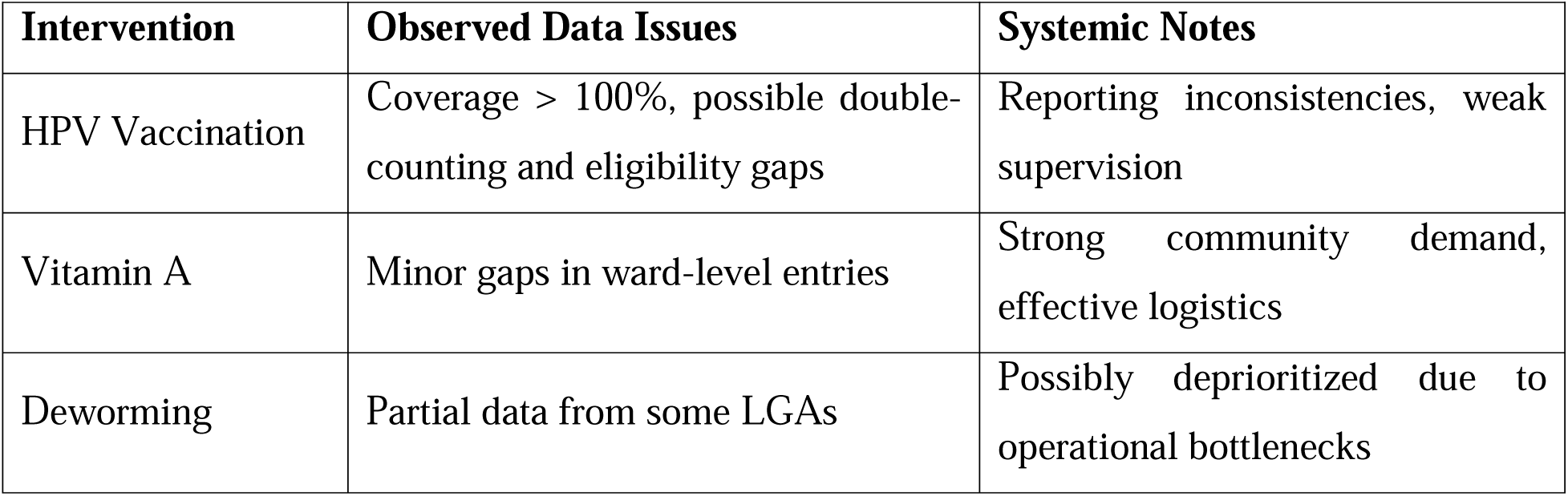
Observed Data Reliability and Systemic Observations by Intervention (OPS Room Reports, 2025)

| Intervention | Observed Data Issues | Systemic Notes |
| --- | --- | --- |
| HPV Vaccination | Coverage > 100%, possible double-counting and eligibility gaps | Reporting inconsistencies, weak supervision |
| Vitamin A | Minor gaps in ward-level entries | Strong community demand, effective logistics |
| Deworming | Partial data from some LGAs | Possibly deprioritized due to operational bottlenecks |

**Table 5:** Identified Operational and Reporting Challenges During MNCH Week (OPS Room Report, 2025)

| Challenge Category | Specific Description |
| --- | --- |
| Delayed Rollout | Southern Ijaw LGA started late due to logistics |
| Incomplete Entries | STF teams failed to upload full service data |
| Missing Reports | Gaps in reporting from Nembe and SILGA |
| Data Tool Deployment Delay | Late release of forms and tools reduced data quality control |
| Dashboard Validation Gap | Mismatch between Power BI and paper-based tallies |

These comparisons provide valuable context, suggesting that while the HPV vaccine may have been prioritized administratively, it may also have been less rigorously monitored in terms of eligibility screening and data quality controls.

### 3.3 Systemic and Operational Challenges in Service Delivery and Reporting

The third objective of this study was to identify and assess the systemic and operational factors that may have influenced the accuracy of the HPV vaccination data. The OPS Room Report revealed multiple process-related weaknesses, particularly in the areas of data entry, reporting timeliness, and service implementation. Notably, Southern Ijaw LGA did not commence MNCH Week activities on the designated Day 1, leading to significant delays in service rollout. Additionally, many STF field officers failed to complete or submit comprehensive reports, especially in hard-to-reach or riverine areas such as SILGA and Nembe. Figure 5 shows that there were also technical lapses in the deployment of data collection tools, which affected the ability of monitoring teams to ensure real-time validation of service delivery. These challenges are outlined in Table 3.3a.

**Figure 5:**
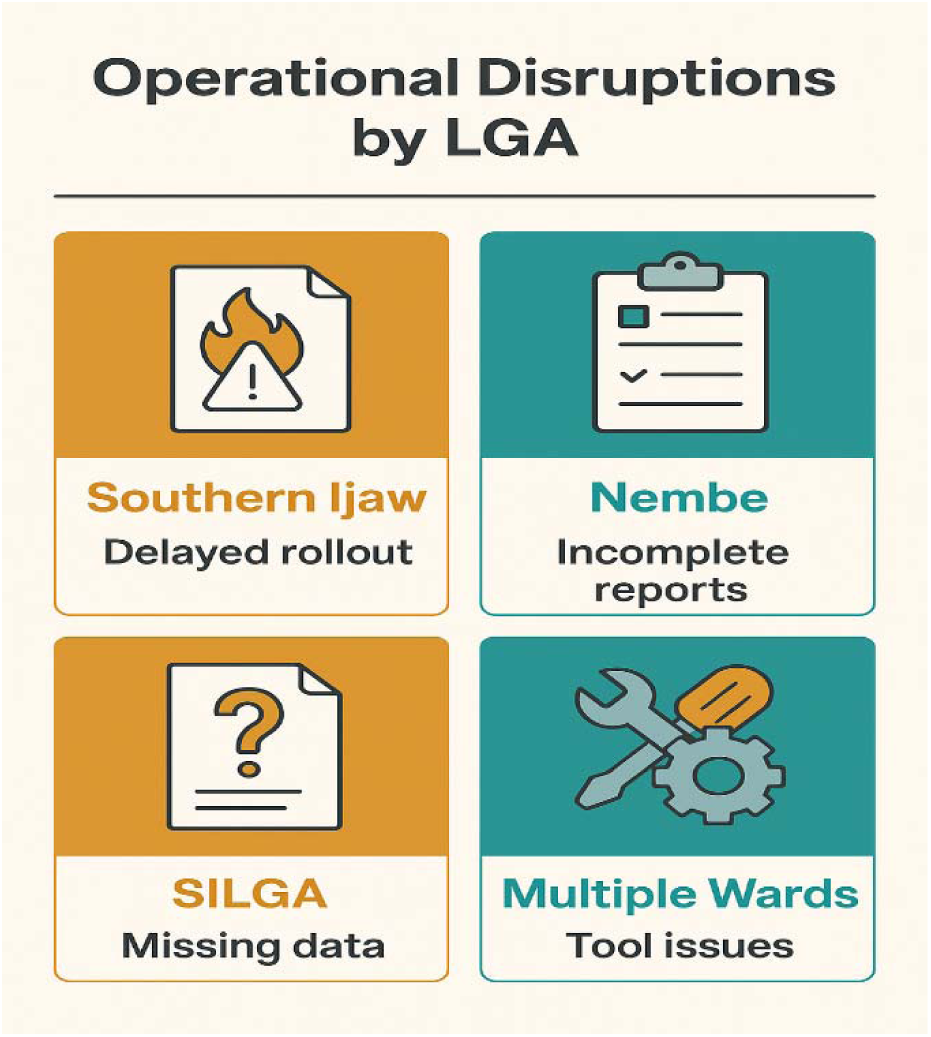
Operational Challenges Across LGAs (OPS Room Report, 2025)

The implications of these challenges were evident across multiple service areas. For example, Routine Immunization (RI) services were not uniformly delivered across all wards, MUAC screening for malnutrition was only partially completed, and the total number of pregnant women who received multiple micronutrient supplements (MMS) was notably underreported in at least two LGAs. These adverse effects are captured in Table 6 and visualized in figure 6.

**Figure 6:**
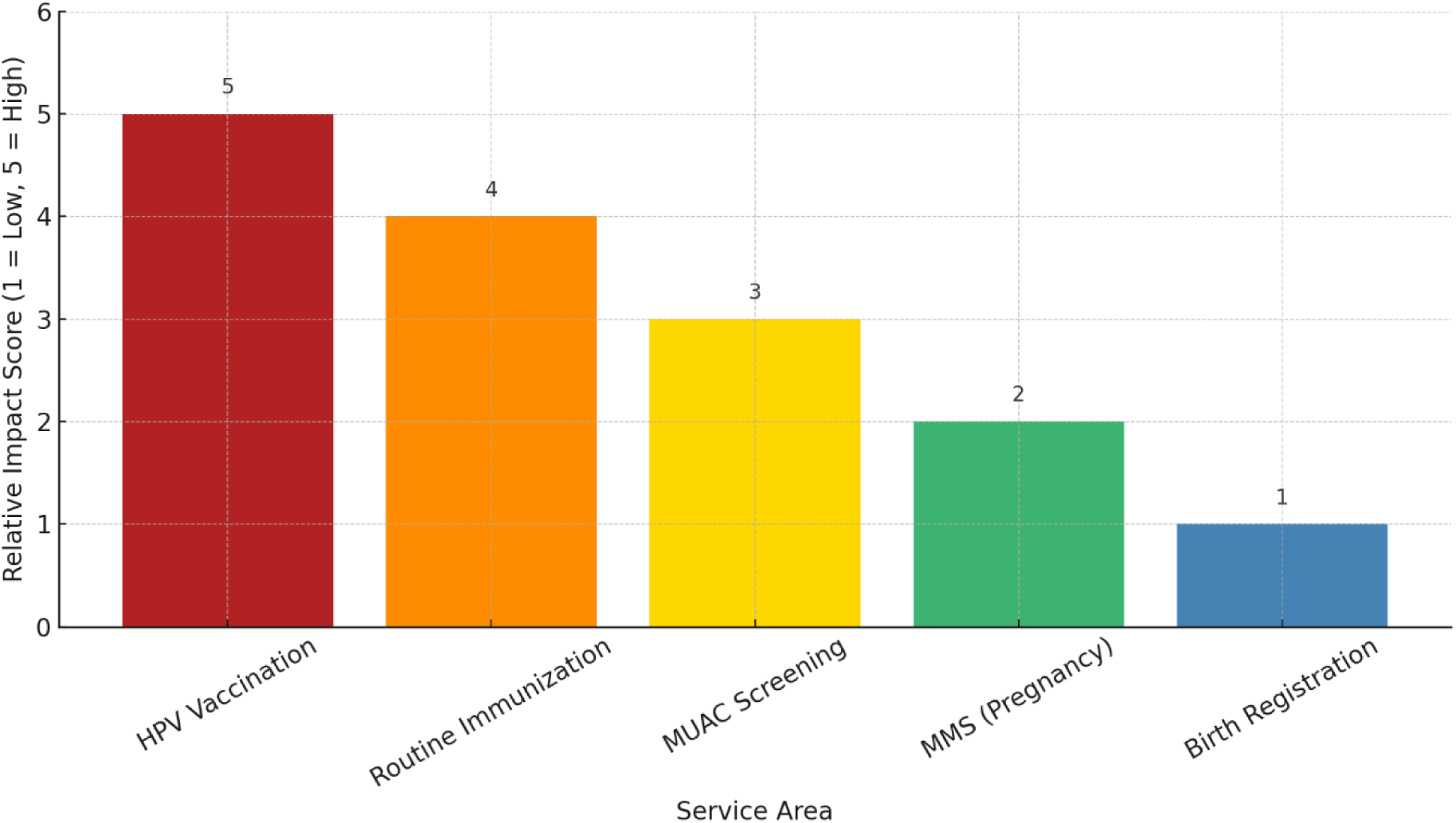
Programmatic Impact of Operational Challenges (OPS Room Report, 2025)

**Table 6:** Programmatic Impact of Operational Challenges Across Interventions (OPS Room Report, 2025)

| Impacted Service | Documented Impact |
| --- | --- |
| HPV Vaccination | Inflated figures, inconsistent tracking |
| Routine Immunization | RI not conducted uniformly across all wards |
| MUAC Screening | Red MUAC identification incomplete or partial |
| MMS (Pregnancy) | Total recipients underreported in 2 LGAs |
| Birth Registration | Only 1% coverage state-wide, very poor data capture |

Overall, these results highlight a complex interplay between logistical inefficiencies, technical delays, and weak supervision, all of which compromised the credibility of reported coverage figures. In particular, the HPV vaccination program, while numerically impressive, appears to have suffered from serious data reliability issues that warrant further investigation and system-level reform.

## 4.0 DISCUSSION

The findings from this study expose critical structural and operational shortcomings in the delivery and reporting of HPV vaccination during the June 2025 MNCH Week in Bayelsa State. While the reported coverage figure of 330% may initially appear as a success story, a deeper examination reveals that such numerical excess signals more problems than progress. When analyzed alongside other MNCH interventions, Vitamin A and Deworming, it becomes evident that the disparities are symptomatic of a fragile delivery system, flawed monitoring, and inadequate governance. These challenges are not unique to Bayelsa but are reflective of broader issues in campaign-based service models across low- and middle-income countries (LMICs).

### 4.1 Inflated Coverage and Data Integrity Concerns

The reported HPV vaccination coverage of 330%, against a target population of 18,122 adolescent girls, suggests that nearly three times more individuals were vaccinated than those originally targeted. This anomaly, rather than indicating commendable outreach, signals a breakdown in eligibility verification, documentation, and data quality assurance. While some level of overshooting may occur due to migration or minor estimation errors, a 230% overreach is implausible under any reliable system.

One probable factor behind this inflation is the absence of standardized age verification procedures at service points. In many campaign settings, health workers lack tools for verifying the age of recipients, relying instead on verbal affirmation or visual estimates, which are prone to error. Furthermore, under pressure to meet output-based targets, some teams may have counted the same individuals more than once, particularly in LGAs with porous geographical boundaries or overlapping outreach teams. In remote or riverine areas, where digital monitoring was delayed or absent, such duplications likely went undetected.

The lack of electronic registers further compounded this issue. In comparison, countries like Rwanda have implemented robust school-based HPV vaccination programs using digital registries that log each dose against a specific student profile. This approach, supported by real-time dashboards, has helped Rwanda achieve sustained coverage rates above 90% with minimal reporting errors. Likewise, Uganda’s pilot programs in HPV vaccine delivery were accompanied by district-level validation mechanisms and age-specific school rosters, which minimized errors and inflated counts. These examples demonstrate that when digital infrastructure and strict eligibility protocols are in place, data reliability significantly improves.

In Bayelsa, however, digital tool deployment was late and inconsistent. Even where Power BI dashboards were available, field officers often lacked the training to input or validate data correctly. This limitation, while acknowledged during campaign debriefs, had not been resolved before final report validation. As a result, inflated figures made it into official summaries, undermining both program credibility and the basis for future planning.

Moreover, the reliance on manual tally sheets without real-time verification mechanisms allowed reporting errors to persist unchallenged. Health workers in remote wards, faced with time constraints, poor supervision, and a lack of feedback loops, likely recorded estimates rather than exact figures. These practices, while understandable in high-pressure contexts, highlight the fragility of campaign-only delivery systems.

### 4.2 Comparative Intervention Gaps and Prioritization Bias

The stark contrast between the 330% HPV coverage and the more realistic 83% and 55% for Vitamin A and Deworming, respectively, highlights not only inconsistencies in implementation but also possible prioritization biases. Vitamin A has long been a staple of MNCH campaigns, with well-established logistics, high community awareness, and consistent funding. Its coverage in Bayelsa, though not optimal, reflects relatively mature delivery systems and a strong history of implementation.

Deworming, by contrast, had the lowest coverage (55%) and suffered from poor mobilization and partial tool deployment in several LGAs. It is possible that health workers, focusing on more “politically visible” or new interventions like HPV, allocated less attention to Deworming. Furthermore, the logistical complexity of administering tablets, the potential for mild side effects, and limited caregiver demand likely contributed to its underperformance.

The HPV vaccine, as a new entrant, should reasonably have experienced start-up challenges, such as hesitancy, awareness gaps, and logistical bottlenecks. Instead, it reported the highest coverage—suggesting that the figures may have been artificially boosted either through overreporting, misclassification of recipients, or weak supervision.

This prioritization bias may also be linked to international donor pressure or national-level incentives to demonstrate early success in HPV rollout. While understandable, such dynamics can distort local decision-making and create incentives for numerical inflation over actual impact. Moreover, since HPV vaccination was implemented without integration into school registers or community enumeration lists, the risk of reaching out-of-scope individuals was high. The absence of proper feedback mechanisms to catch and correct these errors during the campaign further exacerbated the problem.

An important limitation here is that we were unable to access disaggregated data by age, school, or ward, which could have helped confirm the proportion of out-of-scope vaccinations. Without such granularity, it is impossible to determine how many of the 59,803 doses administered were given to truly eligible girls. This lack of disaggregated reporting constrains our ability to assess equity, especially among hard-to-reach or marginalized groups.

Moreover, a missed opportunity exists in aligning HPV vaccination with other adolescent services. Countries like Australia have institutionalized HPV delivery alongside annual health checks and comprehensive school health programs. This not only increases uptake but ensures consistency in data collection, ethical consent, and population coverage validation. Bayelsa’s campaign, by contrast, operated in isolation, without leveraging routine adolescent health infrastructure—thus widening the gap between reported numbers and actual service equity.

### 4.3 Systemic Weaknesses and Operational Fragmentation

Underlying the inflated figures and uneven intervention performance are deeper systemic weaknesses that affect all aspects of campaign planning and execution. Key operational challenges identified in Bayelsa included late campaign rollout in Southern Ijaw, non-submission of complete reports by State Technical Facilitators (STFs) in Nembe and SILGA, and a failure to harmonize manual and digital records. Each of these issues directly contributed to the distortion of final coverage statistics.

Southern Ijaw’s delayed rollout is especially significant. As the largest and most logistically challenged LGA in Bayelsa, its late start likely disrupted the delivery schedule, shortened implementation windows, and forced teams to rush activities. This compressed timeline created conditions where accurate documentation became secondary to meeting numeric targets. Incomplete or hastily filled tally sheets from these areas thus made it into the OPS Room report, contributing to overestimations.

Furthermore, STF teams failed in their role as frontline monitors. Tasked with ensuring data integrity, many were either poorly trained or unsupported during the campaign, especially in water-locked regions. Their incomplete reports meant that data from entire catchment areas were either missing, substituted, or estimated. This not only weakens the confidence in reported coverage but also leaves decision-makers blind to ground-level disparities.

Another significant gap was the inconsistent use of Power BI dashboards. While designed to enhance transparency and real-time validation, their late deployment and inadequate orientation sessions rendered them underutilized. In fact, many LGAs reverted to manual tools due to poor internet connectivity or lack of electricity. These technical limitations meant that mismatches between digital entries and manual summaries went unresolved during the campaign window, allowing inconsistencies to persist into final reports.

Moreover, the campaign lacked a built-in mechanism for second-dose follow-up or post-campaign validation, which are essential for assessing the actual success of HPV interventions. Without tracking whether girls received the correct vaccine dose, on time, and without duplication, the value of the reported figures becomes questionable. Unlike routine immunization—which follows up with health cards, reminder systems, and health facility records—MNCH Week lacks the infrastructure to ensure continuity of care.

The implications of these limitations are profound. Not only do they undermine the credibility of the campaign, but they also create a false sense of security that the target population has been reached and protected. This could lead to premature withdrawal of resources, underfunding of follow-up phases, and persistent vulnerability among the very girls the intervention seeks to protect.

## 5.0 CONCLUSION AND RECOMMENDATIONS

### 5.1 Conclusion

The introduction of the Human Papillomavirus (HPV) vaccine into Nigeria’s biannual MNCH Week campaign structure represents a critical step forward in the nation’s efforts to reduce the burden of cervical cancer and improve adolescent health outcomes. However, the experience documented in Bayelsa State during the June 2025 round demonstrates that numerical success does not always translate into programmatic integrity. While reported coverage stood at an astonishing 330%, this figure must be interrogated rather than celebrated. A close examination of the campaign’s operational and reporting environment reveals a multitude of challenges that significantly dilute the credibility of this outcome and cast doubt on its sustainability.

One of the most striking revelations of the study was the severe disconnect between projected targets and reported outputs. Rather than indicating exceptional outreach, the over-coverage suggests weak verification mechanisms, inappropriate documentation practices, and potential double-counting of recipients. The use of paper-based registers without real-time validation, coupled with a lack of standardized supervision, created an environment ripe for overreporting. Field-level health workers and STF teams operated under significant time pressure and were often inadequately trained to manage a new intervention like the HPV vaccine, which requires strict adherence to age-based eligibility and dosing protocols.

In contrast, more established MNCH services such as Vitamin A supplementation and Deworming demonstrated realistic and historically consistent coverage rates. This differential performance illustrates that the strength of an intervention’s data integrity depends not only on its clinical importance but also on the maturity of its delivery systems. Vitamin A campaigns, for instance, benefit from years of implementation experience, a robust logistics pipeline, and established community demand. In comparison, the novelty of the HPV intervention, its limited public awareness, and logistical uncertainty collectively undermined data consistency and skewed overall outcomes.

Further complicating the situation were the operational challenges encountered across several LGAs. Southern Ijaw, a region particularly affected by access difficulties, experienced delayed rollout, which translated into uneven coverage, confusion among mobilized populations, and misalignment of planning targets. STF teams in other LGAs either failed to report on time or submitted incomplete entries, further eroding confidence in the reliability of the MNCH Week data dashboard. The lack of functional integration between manual entries and Power BI digital systems made it difficult to perform timely verifications, and where mismatches occurred, they were largely unresolved before final reporting.

These experiences reveal a broader structural deficiency in campaign-based health programming. While MNCH Week remains a valuable opportunity to reach vulnerable populations, especially in rural and hard-to-reach areas, it cannot compensate for weak systems and fragmented accountability chains. Campaigns should never be so focused on immediate targets that they ignore data integrity, population equity, or long-term trust-building within the communities they serve. The lesson here is not that the HPV campaign failed entirely, but that its perceived success masks a significant data crisis that must be urgently addressed if future campaigns are to be both credible and impactful.

In essence, this study calls attention to the gap between service delivery and service documentation. It emphasizes the urgent need for stronger governance structures, advanced data verification systems, and more integrated community engagement strategies. By treating data as a public health asset rather than just a compliance requirement, stakeholders can begin to rebuild confidence in the MNCH Week platform, realign it with global immunization goals, and ensure that HPV vaccination efforts contribute meaningfully to reducing cervical cancer risks in Nigeria.

### 5.2 RECOMMENDATIONS

To enhance the credibility, accuracy, and long-term effectiveness of HPV vaccination within Nigeria’s MNCH Week framework, the following ten recommendations are proposed. These are grouped thematically into four strategic clusters:

1. **Introduce Individual-Level Digital ID Capture at Point of Service**: Implement biometric or QR-code–based registration tools linked to national IDs or school records to uniquely track vaccination episodes. This prevents duplication and improves real-time accountability, particularly in high-mobility populations (World Health Organization, 2022; Dixon et al., 2019).
2. **Establish Real-Time LGA-Level Data Reconciliation Protocols**: Set up LGA-based reconciliation hubs where tally sheets are reviewed against Power BI dashboards within 24 hours to ensure early detection of inflation or omissions. Such real-time validation aligns with WHO digital health guidance for immunization systems (Morgan et al., 2022; WHO, 2022).
3. **Integrate HPV-Specific Modules into Mandatory In-Service Training**: Develop a short, certified course for health workers and STF teams on HPV vaccine protocols, adolescent communication, cold chain logistics, and age eligibility. Targeted training significantly improves vaccine documentation accuracy (Deshmukh et al., 2018; Santa Maria et al., 2021).
4. **Develop a Logistics Risk Mapping Tool for Hard-to-Reach Areas**: Design GIS-based tools to identify service bottlenecks in riverine terrain and pre-deploy vaccines and data kits accordingly. Risk-based logistics planning has proven essential for equity in hard-to-reach populations (Elemuwa et al., 2024; Mohamed et al., 2022).
5. **Institutionalize Gender-Sensitive Peer Education Models**: Train adolescent girls as peer educators or “HPV Ambassadors” to demystify the vaccine, reduce stigma, and increase uptake in school and community settings. Peer-led approaches have shown effectiveness in increasing adolescent vaccine acceptance (Chodick et al., 2021; Obulaney et al., 2016).
6. **Launch Mobile Media Vans for Pre-Campaign Sensitization**: Deploy culturally adapted media vans broadcasting in local languages 2–3 weeks before MNCH Week to improve informed consent and parental engagement. Mobile outreach has proven effective in overcoming sociocultural barriers in other vaccine campaigns (Pot et al., 2017; Scarinci et al., 2020).
7. **Make Data Integrity a Performance Indicator for STF Contracts**: Include specific indicators (e.g., data completeness, age-verification compliance, and real-time reporting) in the performance evaluation of STF officers. Linking data quality to contract renewals strengthens accountability (Walling et al., 2016; Dussault et al., 2023).
8. **Create a Cross-Sector HPV Coordination Desk within SPHCDA**: Establish a state-level taskforce involving health, ICT, and education sectors to coordinate HPV activities year-round. Multisector integration has improved vaccination continuity in countries like Australia and Uganda (Davies et al., 2023; Canfell et al., 2015).
9. **Require Public Release of Independent Campaign Scorecards**: Mandate third-party audit reports of MNCH campaign data to be publicly released within 30 days post-campaign, increasing transparency and public trust (UNICEF, 2022; Mohamed et al., 2022).
10. **Transition HPV Delivery to a Hybrid School–Facility Model**: Gradually shift from campaign-only administration toward a hybrid model that combines school-based vaccination with facility-based catch-up days. Countries using this model (e.g., Rwanda, Australia) have achieved sustained and equitable coverage (LaMontagne et al., 2022; WHO, 2023).

### 5.3 Final Thoughts

The expansion of HPV vaccination into Nigeria’s national immunization strategy via the MNCH Week platform offers a vital opportunity to protect adolescent girls from one of the most preventable causes of cancer. It symbolizes the country’s intent to align with global cervical cancer elimination goals and respond to growing advocacy around adolescent health and rights. However, as the Bayelsa case shows, the success of such interventions is not determined solely by the number of doses administered or coverage percentages reported. True success lies in the credibility, inclusiveness, and sustainability of the systems that support service delivery.

What makes HPV vaccination unique is its positioning at the intersection of health service innovation, gender equity, and systems strengthening. Unlike routine childhood immunizations, HPV programs must account for complex social dynamics, including community trust, age sensitivity, and awareness gaps among caregivers and schools. The campaign setting, while useful for rapid outreach, cannot be allowed to mask the structural weaknesses that hinder effective service tracking and population accountability.

Therefore, future rounds of MNCH Week must evolve beyond numerical achievement and move towards value-based public health programming. Every intervention whether it targets a child, a pregnant woman, or an adolescent girl must be backed by systems that ensure data integrity, community ownership, and transparent governance. The lessons from Bayelsa are not isolated; they resonate with similar campaigns across low- and middle-income countries where health systems struggle with fragility, underfunding, and decentralization.

Ultimately, the goal is to create a future where every girl receives the HPV vaccine not because her name appears on a tally sheet, but because the system has reached her, protected her, and accounted for her health needs with accuracy and dignity. This is the standard that health campaigns in Nigeria and similar contexts must now strive to meet. Through reforms grounded in data, equity, and collective accountability, MNCH Week can be transformed from a numerical exercise into a truly people-centered vehicle for public health impact.

## Data Availability

All data produced in the present work are contained in the manuscript

